# Organizational resilience and its implications for healthcare workers in the COVID-19 pandemic: A literature review

**DOI:** 10.1101/2024.10.10.24315244

**Authors:** Karolina Kaczmarski, Atena Pasha, Abdul-Hanan Saani Inusah, Xiaoming Li, Shan Qiao

**Author notes:** **Corresponding author**: Shan Qiao Associate Professor Arnold School of Public Health Department of Health Promotion Education and Behavior SC SmartState Center for Healthcare Quality (CHQ) University of South Carolina Columbia, SC 29082.

## Abstract

**Background:** Organizational resilience is crucial in supporting the well-being of healthcare workers and ensuring the quality of healthcare services during crises like the COVID-19 pandemic. This study aims to comprehensively review organizational resilience of healthcare facilities in terms of its conception, measurement, and impacts on healthcare workers during the COVID-19 pandemic.

**Methods:** A search was conducted in four databases (PubMed, ScienceDirect, PsycINFO, and Web of Science) for empirical articles considering organizational resilience among healthcare facilities during the COVID-19 pandemic from 2019 to 2024. Several keywords from three categories (“COVID-19”, “organizational resilience,” and “healthcare facilities”) were used, and RAYYAN was used to manage references.

**Results:** Four empirical articles from 172 studies were included, which encompassed a total sample of 6,606 healthcare workers from Switzerland, Saudi Arabia, Iran, and Türkiye. Organizational resilience could influence the individual resilience of healthcare practitioners, enhance crisis management and ensure safety performance. The strategies of enhancing organizational resilience at healthcare worker level included staff training, crisis management protocols, collaboration promotion, and stress management approaches. The ones at health facility level included government intervention, funds for hospital preparedness, competency-based crisis management, and mental health programs for healthcare workers. Our review also suggests a lack of empirical studies, no commonly used measurement instruments, and the heterogeneity of study contexts in the research of organizational resilience in public health.

**Conclusions:** This review highlights effective strategies to enhance the organizational resilience of healthcare workers and examines their impact during the COVID-19 pandemic. Immediate government action, funding to support hospital preparedness, and the formation of flexible healthcare teams are essential to strengthen organizational resilience among healthcare workers for future crises.

## INTRODUCTION

The COVID-19 pandemic has resulted in substantial social and lifestyle changes and significantly impacted both physical and global mental health. The impact of COVID-19 on healthcare workers has been particularly profound, as they have been exposed to exceptionally stressful conditions compared to many other professional groups. As COVID-19 cases surged and patient numbers grew, healthcare workers faced significant mental and physical health challenges, intense stress, heavy workloads, burnout, and the risk of moral injury(Greenberg et al., 2020; Heath et al., 2020; Jalili et al., 2021; Krystal, 2020; Sasangohar et al., 2020). Working under such stress can lead to declines in job performance and adverse effects on both social and familial aspects of life, such as increased infection risks, fear of spreading the virus to others, social isolation from loved ones, longer work hours, and challenging work environments with extended shifts(Conway et al., 2008; Manchia et al., 2022). This stress can have negative impacts on well-being, psychological and behavioral factors, task performance, cognitive function, and safety(Moradi et al., 2014; Omidi et al., 2018; Park & Kim, 2013). According to (Elliott et al., 2021), these circumstances have directly affected the mental health of healthcare workers, leading to a 46% increase in depression rates and a 30.1% increase in anxiety rates following quarantine. It is likely that these rates are even higher for nurses and healthcare workers who have direct, regular contact with patients compared to physicians and other healthcare professionals.

Research indicates that organizational resilience, characterized by the ability to anticipate potential threats, effectively manage challenging situations, and adapt to changing circumstances and unexpected disruptions, is associated with positive outcomes and institutional success (Salehi et al., 2023). Additionally, organizational resilience has been shown to improve psychological well-being and address the daily occupational demands of individuals (Brown, 2021). Prior studies also underscore the importance of considering organizational resilience in protecting the mental health of nurses and healthcare professionals (Rivas et al., 2021; Smith et al., 2023).

The focus on organizational resilience and the adaptive response of health services and hospitals during the COVID-19 pandemic will promote the development of new capacities to address not only the current pandemic but also future emergent situations. By fostering organizational resilience among healthcare facilities, we can cultivate confident professionals with strong self-reflection skills, analytical thinking abilities, and adaptability in the face of unprecedented challenges. Previous studies can serve as inspiration to further investigate how increased organizational resilience among health facilities can not only decrease the academic gap (caused by limited empirical data and varied pandemic approaches) but also provide practical implications to the healthcare system (Manchia et al., 2022).

However, there is a lack of research comprehensively considering organizational resilience, its associated factors, measurement instruments, impacts, and strategies during the COVID-19 pandemic among healthcare workers. Previous systematic reviews have focused on the effects of the pandemic on overall mental health outcomes among healthcare workers instead of particularly investigated resilience (Chemali et al., 2022; De Pablo et al., 2020; Neto & Silva, 2023; Sheraton et al., 2020). A few studies explored resilience, but most of them reviewed resilience as a concept at the individual level (Baskin & Bartlett, 2021; Heath et al., 2020; Rieckert et al., 2021). Only one study considered organizational resilience in the health system context focusing on measuring the resilience of healthcare facilities (Ignatowicz et al., 2023).

To address these knowledge gaps, the current study aims to explore organizational resilience and its impacts on healthcare workers during the pandemic and to identify strategies of improving resilience within healthcare settings. Specifically, the study aims to focus on 1) the conceptualization of organizational resilience among healthcare facilities and its measurement instruments; 2) the impacts of organizational resilience on healthcare workers in terms of the psychosocial well-being of healthcare workers and the quality of care provided during the COVID-19 pandemic; and 3) the effective strategies to enhancing organizational resilience among healthcare facilities.

## METHODS

### Search Strategies and Data Sources

A comprehensive search was conducted in September 2023 across Electronic Bibliographic Databases (PubMed, PsycINFO, ScienceDirect, and Web of Science), Reference Lists of Reviews and Meta-Analyses, and Reference Lists of Included Studies. Table 1 outlines the databases according to name, topics covered, and frequency of updates for each catalog. The search strategy included keywords from three categories: COVID-19 (e.g., COVID-19), organizational resilience (“organizational resilience”), and healthcare workers (e.g., Health Personnel, Health Facilities). Keywords can be found in Table 2 of the supplementary document.

**Table 1.** Description of Records.

| Author/s, year | Country | Target Population | Organizational Resilience Definition | Organizational Resilience Measurement | Strategies to enhance organizational resilience in healthcare facilities | Key findings |
| --- | --- | --- | --- | --- | --- | --- |
| *Balay-Odao et al. (2021)* | Saudi Arabia | Clinical nurses | Ability to face adverse situations while remaining focused and optimistic for the future | Hospital Preparedness Assessment Tool was used to measure hospital preparedness, as a factor of organizational resilience. | **Anticipatory or preventive measures**<br>Enhance continuing education, train staff on infection control, rapid case identification, use of PPE<br>**Appropriate management during the crisis**<br>Follow guidelines, coordinate with local authorities, isolate suspected COVID-19 patients, maintain hygiene, restrict visitor access.<br>**Psychological support**<br>Provide peer support, encourage spiritual practice, boost positive emotions (confidence, calm), organize mental health programs, sufficient government funding. | **COVID-19 hospital preparedness and resilience** Nurses perceived high preparedness, facilitated by swift government action and resources (e.g., funds, medical supplies and equipment)<br>Resilience increased with increasing age and experience, enhancing problem-solving skills and overall resilience.<br>Confidence in public health authorities improved nurse confidence in handling COVID-19 cases. Competent nurses coped better with stress, leading to stable mental health levels.<br>**Psychological burden**<br>Nurses caring for COVID-19 patients had lower anxiety scores.<br>Male Saudi nurses as well as Christian, and Hindu nurses were more vulnerable to psychological stress burden, but high resilience helped mitigate depression risks. |
| *Juvet et al. (2021)* | Switzerland | All the staff of a university hospital in French-speaking Switzerland: | Processes and capacities that enable individuals or systems to overcome, resist, adapt to | Utilized two qualitative questions to assess organizational resilience | **Anticipatory or preventive measures**<br>Improve adaptive capacity, provide skill training, foster interdisciplinary collaboration, and improve communication<br>**Appropriate management during the crisis** | Of 4,773 participants, 1,292 reported challenges such as organizational changes (41.8%), task and logistics reorganization (17.3%), adapting to new roles (15.6%), and modified patient flows (7.7%). |
|  |  | healthcare staff, support staff, frontline staff and administrative staff | and recover from deep crises. |  | Implement self-regulation strategies promote teamwork, reorganize tasks and spaces, ensure accurate information dissemination, maintain adequate PPE stocks, prioritize interprofessional collaboration, and adopt a systemic safety.<br>**Psychological support**<br>Offer peer and patient support, engage in multidisciplinary discussions, balance situational challenges with values, manage contamination fears and reorganize family life. | Changing dynamics due to new colleagues were noted (7%), and the need for quick adaptation was common (5.2%).<br>Surgical departments reported more problematic changes (59.2%) compared to non-hospital institutions (34.8%).<br>Workload issues, including overloads (9.8%) and irregular schedules (9.6%), were prevalent.<br>Under-staffing led to significant challenges (7.4%), while lack of breaks and work-life balance issues were also highlighted (5.4%, 0.9%)<br>Concerns about patient care quality were noted, especially among healthcare workers under age 40 (20.7%) compared to older workers.<br>Isolation from families due to visiting restrictions (5.2%) was also problematic. Employees in COVID-19 wards reported more quality care issues (23.9%) than those in non-COVID wards (16.2%). |
| *Salehi et al. (2023)* | Iran | Healthcare providers employed at a teaching hospital | An organization's ability to anticipate potential threats, | Sixteen items from Hollnagel (2013) resilience tool. | **Anticipatory or preventive measures**<br>Build reserves pre-crisis, foster a supportive workplace culture and prioritize effective leadership.<br>**Appropriate management during the crisis** | For healthcare providers aged 41–50 and over 50 organizational resilience significantly influenced safety performance with weights of 42% and 45%, respectively.<br>Experienced providers (15-20 years) and those with over 20 years reported organizational |

| Author/s,<br>year | Country | Target<br>Population | Organization<br>al Resilience<br>Definition | Organizational<br>Resilience<br>Measurement | Strategies to enhance organizational<br>resilience in healthcare facilities | Key findings |
| --- | --- | --- | --- | --- | --- | --- |
|  |  |  | manage adverse events effectively, and adapt to changing conditions and sudden disruptions |  | Managers should consider the specific needs and resources of employees of different ages.<br>**Psychological support**<br>Implement stress management strategies to reduce occupational stress. | resilience as a more critical factor than others, with weights of 47% and 56%, respectively.<br>Occupational stress notably has affected women's safety performance (31%). |
| *Yağmur and Myrvang (2023)* | Türkiye | Healthcare professionals working in different positions in public health institutions | The capacity to survive, adapt, and grow in the face of changes maintenance of a positive adjustment under challenging conditions such that the organization emerges from those conditions strengthened and more resourceful | Organizational Resilience Scale | **Anticipatory or preventive measures**<br>Focus on equipping managers and personnel with knowledge and experience, adopting a competency-based crisis management approach, enhancing organizational responsiveness, and improving organizational agility<br>**Appropriate management during the crisis**<br>Develop flexible organizational structures, create expert teams, conduct trainings and seminars, and implement regularly updated crisis management plans | Organizational resilience has a positive correlation with competency(r:0.519), flexibility (r:0.443), responsiveness (r:0.502), speed (r:0.511), organizational agility (r:0.566) and crisis management (r: 0.671). |

Keywords were organized using the following logical operators: (1) keywords within one category were lined using the OR operator (e.g., SARS Coronavirus 2 Infection OR 2019-nCoV Infections), and (2) keywords across different categories were connected using the AND operator (e.g., SARS Coronavirus 2 Infection AND Organizational resilience AND Health Personnel).

### Eligibility Criteria

The study’s inclusion criteria encompass research examining organizational resilience among healthcare workers during the COVID-19 pandemic, utilizing empirical research methods, and published between 2020 and 2023 in English. Additionally, the studies must involve human samples. The exclusion criteria comprise papers that do not align with the convergence of organizational resilience in healthcare workers during the COVID-19 pandemic, non-peer-reviewed articles (such as editorials, comments, reports, review articles, dissertations, theses, and books), studies presenting only qualitative data, published before 2020, in languages other than English, involving organisms other than humans (such as animal experiments), and purely medical, biological, or pharmaceutical papers.

### Data screening

Articles were initially retrieved and screened based on the eligibility criteria outlined above using the data management software, Rayyan. The study selection process consisted of seven phases (Figure 1). First, all the duplicate studies were removed. Second, two researchers (KK, AP) screened all titles and abstracts. Studies that did not consider organizational resilience among healthcare workers during the COVID-19 Pandemic were removed. Third, two researchers (KK, AP) removed studies that were not relevant to the empirical research method or were written in a language other than English. Fourth, two researchers (KK, AP) conducted full-text screening of the remaining articles and removed studies that were nonpeer review article, or were pure Medical, Biological, and pharmaceutical papers. Fifth, studies that presented only qualitative data were removed. Sixth, studies published before 2020, were removed. Seventh, two researchers (KK, AP) removed studies that were conducted on non-human organisms. Finally, articles that met the inclusion criteria remained and were selected. Any discrepancies were discussed between AP and KK.

**Figure 1.**
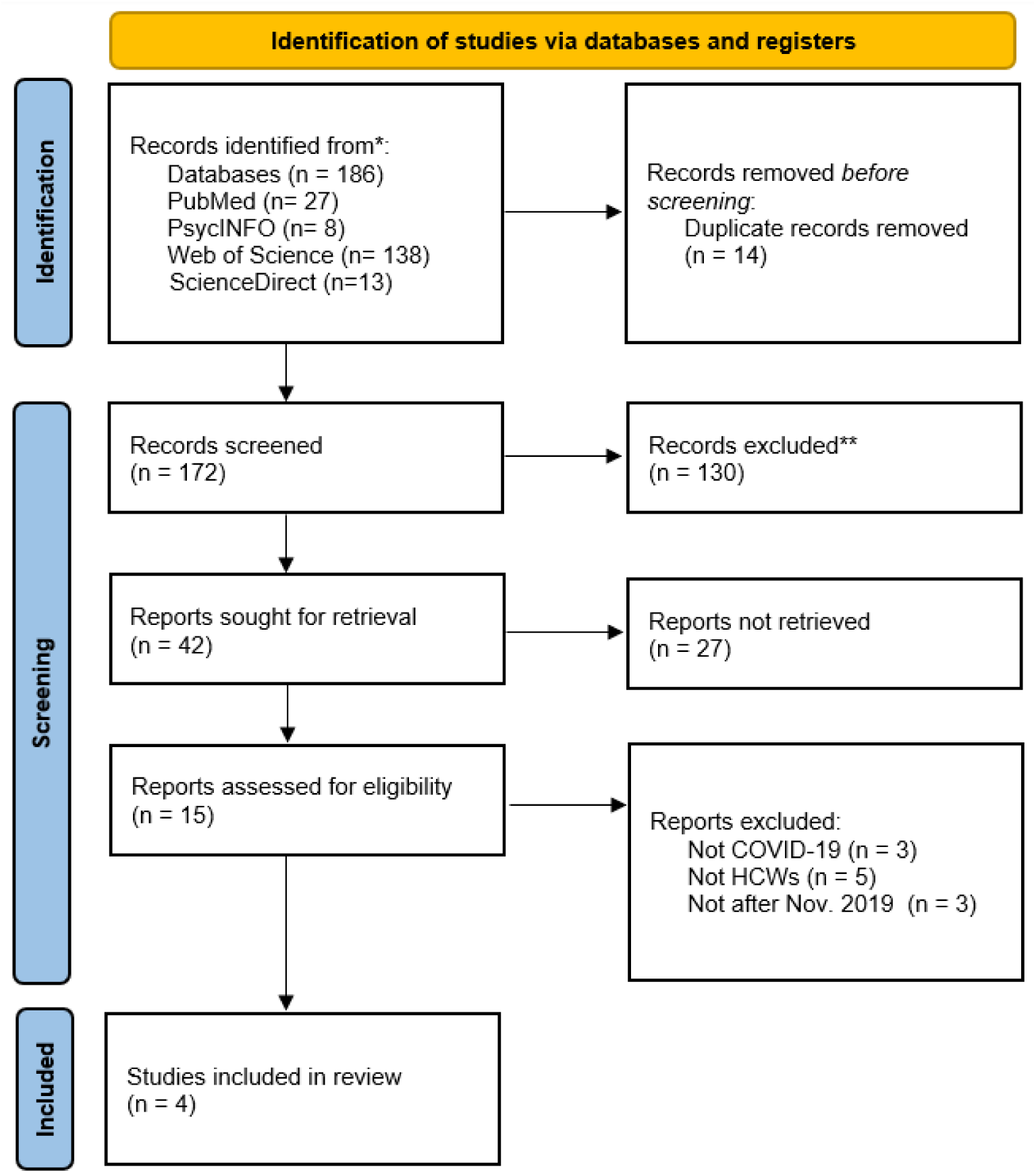
PRISMA 2020 flow diagram from the Electronic Bibliographic Database Searches.

### Data extraction

The data extraction process involved two reviewers using Microsoft Excel. Two forms were created for capturing information from the selected studies. The first form gathered general information, such as citation, title, abstract, keywords, journal, publisher, Doi, year of publication, country, sample size, target population, organizational resilience definition, purpose and process, condition of healthcare providers, COVID-19 stage, intervention, outcomes, limitations, and future suggestions. The second form focused on extracting information related to organizational resilience measurement, including the name of the measurement, format, study design and methods, dimensions, internal consistency, and scoring system. The characteristics of the finally included papers were presented in Table 1.

### Data Analysis

The data analysis process involved employing both qualitative and quantitative methods to integrate findings from the included studies. Descriptive statistics were used to outline study characteristics such as geographical distribution, sample sizes, and key demographics of the involved healthcare workers. Through thematic analysis, common themes and approaches for enhancing organizational resilience were identified by categorizing and grouping similar concepts to form overarching themes. These themes encompassed strategies at both the healthcare worker and health facility levels, addressing factors like staff training, stress management, government intervention, and crisis management protocols. To gauge the impact of organizational resilience on healthcare workers, we examined reported outcomes related to their psychosocial well-being, job performance, and safety, evaluating both direct and indirect effects of resilience strategies on these outcomes. By combining findings from the thematic analysis and impact assessment, we presented a comprehensive overview of effective strategies for bolstering organizational resilience. Furthermore, our synthesis highlighted gaps in the existing literature and suggested potential areas for future research.

## RESULTS

### Description of articles included in the review

We identified four articles that conceptualized organizational resilience as a quantifiable exposure. Using data from a self-report questionnaire, Balay-Odao et al., (2021) administered a cross-sectional design in two government hospitals in Riyadh Region, Saudi Arabia, to determine predictors of hospital preparedness in managing COVID-19 patients and the psychological burden and resilience among clinical nurses. Juvet et al., (2021) conducted a qualitative evaluation using a self-administered online questionnaire adapted from the Working Conditions and Control Questionnaire to determine what problematic situations healthcare employees faced in their daily lives during the first wave of the pandemic, as well as what resilience strategies the individual staff, teams, and institutions implemented to anticipate and adapt to these situations. Furthermore, Salehi et al., (2023) analyzed questionnaires using the entropy and multi-criteria decision-making (MCDM) methods to identify the effect of occupational stress and resilience-related factors on healthcare providers’ safety performance during the COVID-19 pandemic. Finally, Yağmur & Myrvang, (2023), through correlational research design, collected data using the Personal Information Form, Organizational Agility Scale, Crisis Management Scale, and Organizational Resilience Scale to determine the organizational agility levels in health organizations and determine the effect of agility on crisis management process practices and organizational resilience.

### Conceptualization of organizational resilience

Organizational resilience was defined as either an ability or a process. Resilience is defined as an ability, such as the ability to face adverse situations (Balay-Odao et al., 2021), the ability to remain stable during and after crises (Juvet et al., 2021), ability to anticipate and withstand potential threats (Salehi et al., 2023) and the ability to withstand stress (Yağmur & Myrvang, 2023). Resilience is also characterized as a process, such as the processes and capacities to overcome and recover from crises (Juvet et al., 2021). Two levels of organizational resilience were highlighted, individual level and institutional/facility level. The definitions by Balay-Odao et al., (2021) and Salehi et al., 2023 describe resilience at the individual level, particularly for healthcare workers and individuals within organizations. Juvet et al., 2021 and Yağmur & Myrvang, (2023) focus on resilience at the healthcare system level, highlighting the capacity of systems and organizations to adapt and recover from crises.

Organizational resilience is a multifaceted concept described across various studies. All four definitions emphasize the ability to adapt and recover from unforeseen and adverse crises. Juvet et al., (2021); Salehi et al., (2023); Yağmur & Myrvang, (2023) also highlighted stability and functionality as components of organizational resilience. Balay-Odao et al., (2021) further defined organization resilience as the ability to maintain focus and optimism for the future. In addition of anticipating and responding to potential threats, Salehi et al., (2023) also characterized organizational resilience as the ability of organizations to improve the overall safety of a system. The common domains of organizational resilience include anticipation (preparing for potential threats), monitoring (assessing and anticipating potential threats), response (effectively mitigating and managing crises), and learning (adapting and improving based on experiences).

### Measurements to quantify organizational resilience

Balay-Odao et al., (2021) examined the readiness and resilience of both Saudi and non-Saudi clinical nurses in coping with the challenges of the pandemic. They utilized the Hospital Preparedness Assessment to gather data. The Hospital Preparedness Assessment Tool, adapted from the CDC protocol, evaluates a facility’s infection prevention and control policies, focusing on rapid patient identification and isolation, patient placement, transmission-based precautions, hand hygiene, environmental cleaning, HCP monitoring, and visitor access management to ensure readiness in handling confirmed or suspected COVID-19 cases.

To assess organizational resilience, Juvet et al., (2021) utilized two open-ended questions to explore anticipatory and adaptive measures that were implemented by healthcare teams in response to a problematic situation encountered during the COVID-19 pandemic. Answers to these two questions were presented together and categorized into four main themes of resilience: organizational, team, equipment, and individual.

Salehi et al., (2023) used sixteen items prepared by Hollnagel, (2013) to assess the influence of organizational resilience factors on healthcare providers’ safety performance. This measure considers the four abilities of organization resilience: response, monitoring, anticipation and learning. For example, an item assessing the response dimension is: “Does the hospital have the ability to respond to unexpected situations, such as increased number of patients or emergencies”, The monitoring dimension includes items like “Have specific indicators been defined to monitor the current state of the hospital?”. An example of a learning item is: “Is there formal and continuous organizational training for learning in the hospital”. Finally, the anticipation dimension includes items such as, “Has the hospital’s management established a clear framework outlining the roles and responsibilities of nurses in responding to future incidents and emergencies?”.

Yağmur & Myrvang, (2023) used the Organizational Resilience Scale in their study. This measurement instrument was initially developed by Wicker et al., (2013) and later adapted into Turkish by Çoban Kumbalı, (2018). It is composed of four sub-dimensions—robustness, redundancy, resourcefulness, and rapidity. An example item for robustness dimension is: “Our organization can withstand stress without losing focus.” The redundancy dimension includes the item, “Our organization can employ alternative options to sustain operations during unexpected events,” the resourcefulness dimension includes the item, “Our organization can mobilize resources during unexpected events” and, finally, the rapidity dimension includes the item, “Our organization can adapt quickly to changing circumstances.”

### Factors related to organizational resilience

Balay-Odao et al., (2021) used multiple regression analysis to identify factors influencing nurses’ resilience during the COVID-19 crisis. The analysis revealed that age, years of experience, nurses’ area of assignment, confidence levels in public health authorities and hospital administration, and the hand hygiene subscale of hospital preparedness were significant predictors. Specifically, older age and more years of experience were associated with higher resilience levels, while increased confidence in public health authorities correlated with greater resilience. Conversely, heightened confidence in the government and hospital administration was linked to lower resilience levels.

According to Juvet et al., (2021), organizational resilience in healthcare is shaped by a range of structural, relational, and individual factors. These include resource (i.e. equipment) availability, preparedness and planning, effective information management, governance processes, leadership practices, organizational culture, and human capital. Social networks and collaborative efforts are also essential. Their analysis suggested that frontline staff’s practical experiences contribute to organizational resilience.

Yağmur & Myrvang, 2023 demonstrates that organizational resilience can be strongly influenced (β = 0.643) by organizational agility (measured by four domains including competency, flexibility, responsiveness, and speed of response). That is, if an organization’s competency, flexibility, responsiveness and speed of response are improved, their organizational resilience will also be enhanced. Their study also found a positive correlation between crisis management and organizational resilience, showing that organizations that are good at crisis management are also more resilient, and vice versa.

### Impact of organizational resilience on mental health and quality of care in the facilities

Balay-Odao et al., 2021 indicated that heightened levels of resilience among nurses were linked to decreased depression scores. Furthermore, heightened trust in public health authorities and nursing administration correlated with lower anxiety scores, while increased confidence in hospital administration was associated with elevated anxiety scores.

Juvet et al., 2021 noted that while frontline healthcare workers, especially caregivers and doctors faced mental health issues due to decision-making in times of uncertainty, anticipatory strategies that strengthened organizational resilience allayed the fears and improved the mental wellbeing of healthcare workers. This study also revealed that healthcare workers expressed emotional burdens associated with several problematic situations, including organizational changes and workloads. These mental health burdens however could be managed by collective and individual measures (peer support, management support and individual management of fear).

Salehi et al., 2023 conducted a study at a teaching hospital in Iran, recognized as a referral center for COVID-19 cases. They surveyed 344 healthcare professionals across various roles who interacted directly with COVID-19 patients. The study suggested that the impact of organizational resilience on safety performance of healthcare providers moderated by age and work experiences. Organizational resilience was found to be the most influential factor affecting the safety performance in age groups 41–50 years and those aged over 50. Similarly, for healthcare providers with over 15 years of work experience, organizational resilience had a significant impact, with a weight of 47%, and for those with more than 20 years of experience, this factor was even more influential, with a weight of 56%.

### Strategies to improve organizational resilience in health facilities

#### Anticipatory or preventive measures

To promote organizational resilience and prepare well for future public health emergencies, it is essential to improve continuing education programs, training healthcare personnel on infection prevention and control policies, rapid identification, as well as transmission-based precaution using appropriate personal protective equipment (PPE) (Balay-Odao et al., 2021; Juvet et al., 2021). At the organizational level, adaptive capacity, interdisciplinary collaboration and workplace cultures, and effective communication were highlighted to ensure the managers and personnel were knowledgeable and experienced, adopting a competency-based approach for successful crisis management, bolstering the capacity of organizations to respond to and handle crises, and improving organizational agility (Salehi et al., 2023; Yağmur & Myrvang, 2023).

#### Appropriate management during the crisis

One of the top priorities for crisis management during the COVID-19 pandemic focused on infection prevention and control within clinical practices, for example, the implementation of proper guidelines and procedures within facilities in collaboration with local authorities, appropriate isolation of suspected or confirmed COVID-19 patients, moving patients with confirmed or suspected COVID-19 within the facility, hand hygiene, environment cleaning, and limiting visitor access (Balay-Odao et al., 2021). It is also recommended to employ a more systemic approach to safety and risk prevention to improve risk awareness, enhance PPE access, and reinforce coping strategies (Juvet et al., 2021). Some studies emphasized the integration of individual and organizational needs in crisis management by using bottom-up strategies, including increasing employee versatility, prioritizing tasks, interprofessional collaboration, cooperation within networks, peer support, and effective communication (Juvet et al., 2021). Healthcare workers were encouraged to apply self-regulation strategies to balance demands and resources as well as personal and professional lives (Juvet et al., 2021). The specific needs and resources of employees of different ages should be sufficiently considered (Salehi et al., 2023). The efforts to improve crisis management should be aligned with the organizational level capacity building. The recommendation strategies include the construction of flexible and adaptable organizational structures, the creation of expert teams, and the organization of seminars, training, conferences, and teamwork activities. The improvement of organizational resources and governance is needed. Healthcare facilities should also regularly update action plans and crisis management procedures for effective responses to future public health crises (Yağmur & Myrvang, 2023).

#### Psychological support for healthcare workers

All the included studies highlighted the importance of enhancing the psychological well-being of healthcare workers during a public health emergency to increase the organizational resilience of healthcare facilities, which, in turn, could provide a positive and supportive environment for healthcare workers. Although the specific strategies should be tailored to various clinical settings, measures recommended by the existing studies include 1) stress management approaches to reduce job burnout and occupational stress through managing fears of contamination, uncertainty, and the unexpected, finding a balance between situational constraints and values, and family-life reorganizations(Juvet et al., 2021; Salehi et al., 2023); 2) social support within facilities, such as peer support from colleagues, multidisciplinary discussions and formation of mixed teams with various expertise, task and emotional support (Balay-Odao et al., 2021; Juvet et al., 2021) and 3) mental health program to promote resilience among healthcare workers. For example, health facilities could allow staff to freely practice their spirituality, facilitate the strengthening of positive emotions such as self-confidence, calmness, relaxation, and cheerfulness, and organize mental health programs to facilitate resilience (Balay-Odao et al., 2021).

## DISCUSSION

This literature review provides a comprehensive overview of organizational resilience, including its conceptualization, correlates, impacts on healthcare workers’ mental health and their quality of care, as well as the strategies to improve organizational resilience during the COVID-19 pandemic. Our findings suggest that organizational resilience had an evident impact on the mental health of healthcare workers and their quality of care during the pandemic. Resilience in healthcare facilities reduced healthcare workers’ stress and burnout and improved patient safety and care quality. Age and work experiences may moderate the impacts of organizational resilience on the quality of care. For instance, the association between organizational resilience and safety of care was more significant among older healthcare workers and those with more work experience. To improve organizational resilience, facilities should invest in leadership training, establish effective crisis management protocols, promote mental health programs, and secure adequate government funding to ensure preparedness for future crises.

Our review also shows several knowledge gaps regarding the organizational resilience of healthcare facilities during public health emergencies. First, existing organizational resilience measurement tools are inconsistent and often lack grounding in theoretical frameworks, making it difficult to establish standardized assessments across different healthcare settings. Balay-Odao et al., (2021) used the Hospital Preparedness Assessment Tool, as measure for organizational resilience. Juvet et al., 2021 made two open-ended questions to explore the anticipatory and adaptive measures implemented by healthcare institutions in response to challenges brough on by COVID-19. Salehi et al., 2023 utilized sixteen items from (Hollnagel, 2013) resilience tool. Yağmur & Myrvang, 2023 used the Organizational Resilience Scale. The inconsistent and inexplicit measurements of organizational resilience reflect a lack of commonly agreed conceptualization regarding organizational resilience. More theoretical research is needed to develop operational concepts about organizational resilience in the context of public health. The solid theoretical framework and widely accepted definition will be the basis on which we can capture the multidimensional and nuanced nature of organizational resilience.

Second, while factors associated with organizational resilience were identified at various socio-ecological levels, gaps exist in addressing all levels comprehensively, with certain areas, including community and policy-level influences, which remained underexplored. Existing literature has identified individual-level, interpersonal-level, and institutional -level factors, including frontline staff’s practical experiences (e.g., their problem-solving skills during crises, familiarity with operational workflows, and awareness of potential risks)(Juvet et al., 2021; Sutcliffe, 2003), social support from colleagues and leadership (Juvet et al., 2021; Shanafelt et al., 2020; Sull et al., 2015), effective leadership, confidence and trust in public health authorities (Akgün et al., 2023; Balay-Odao et al., 2021), and organizational agility and crisis management (Duchek, 2020; Yağmur & Myrvang, 2023). However, there is limited evidence to explore how community-level or policy-level factors may affect organizational resilience. Community-level factors, such as public trust, community engagement in crisis preparedness, and local resources, alongside policy-level factors like government funding and policies, form critical external support outside healthcare institutions (Biddle et al., 2020; Bishai et al., 2024). These factors shape how healthcare facilities prepare for and respond to crises, ensuring essential support for healthcare providers during emergencies. Resilience is not only built within the organizations but is largely influenced by the broader context in which healthcare institutions operate. Addressing these gaps can enhance our understanding of organizational resilience and improve preparedness for future public health emergencies.

Lastly, there is a shortage of intervention studies to test and support policies or recommendations, leaving a gap in evidence-based strategies to effectively enhance organizational resilience in healthcare facilities. Extant literature suggests multiple strategies and recommendations to promote organizational resilience among healthcare facilities, including promoting the spiritual and psychosocial well-being of healthcare workers during the crisis, providing social support, implementing mental health programs, developing effective crisis management strategies, and capacity building to promote organizational agility and adaptivity. However, there is a dearth of specific interventions targeting the organizational resilience of healthcare facilities because of a lack of solid theoretical framework and the difficulties in measurement and intervention evaluations.

Despite these limitations, existing literature can contribute to informing effective preparedness strategies for future public health emergencies. First of all, building up organizational resilience among healthcare facilities is a promising approach to promoting mental health conditions and enhancing their quality of care during the crisis. Empirical studies suggest organizational resilience may reduce depression level, decrease fears due to uncertainties, and improves the psychological well-being (Brown, 2021; Carmassi et al., 2020). Organizational resilience featured by sufficient training, effective preparedness, timely communication, and solid support can be a protective factor against mental health challenges such as stress, anxiety, and burnout for healthcare workers in high-stress environments like healthcare facilities during pandemics (Brooks et al., 2018; Maslach et al., 2001).

Second, interventions to promote organizational resilience should be tailored to specific clinical settings, especially to adapt to the characteristics of healthcare workers within the facilities. Healthcare workers’ age and years of experience were significantly associated with organizational resilience. Older and more experienced healthcare workers were found to consistently demonstrate higher levels of resilience. For example, experienced healthcare workers demonstrated greater resilience in high-stress environments (Kim et al., 2023). Hart et al., (2014) found that experience contributed to enhanced coping mechanisms and psychological well-being among nurses during the pandemic. These studies suggest that individual attributes such as age and professional experience are critical for resilience due to the accumulation of knowledge, coping strategies, and professional confidence over time. Therefore, organizational resilience interventions may particularly pay attention to young healthcare workers who need more support and guidance in coping with unexpected events and uncertainties during the pandemic; meanwhile, leveraging the intensive experiences of older healthcare workers and empowering them as role models could be critical for a successful intervention.

Last but not least, existing studies suggest that organizational resilience enhancement should be integrated into the continuous efforts of capacity-building at the individual-, interpersonal-, and institute-level covering all stages from pre-, during-, and post-crisis stages. Continuous training for healthcare workers, a positive work environment, and reliable social support from interdisciplinary teams in daily practice will facilitate organizational resilience during public health emergencies. Healthcare facilities with higher agility, characterized by rapid decision-making and flexibility in operations, were better able to adapt to the rapidly changing demands during the pandemic, thereby enhancing their overall resilience. At the pre-crisis stage, risk reduction, including anticipation of future crises and actions of preventive measures is a cost-effective strategy to enhance organizational resilience. Proactive risk management strategies are essential for reducing vulnerabilities before they escalate into crises (Lengnick-Hall et al., 2011). For example, by investing in capacity building and appropriate infrastructure, organizations can minimize the potential negative impacts of future emergencies. These measures not only improve safety and reduce costs but also enhance the long-term sustainability and reputation of healthcare institutions. At the stages during and after the crisis, appropriate crisis management strategies are key in terms of mitigating negative impacts and facilitating recovery (Carley et al., 2020). As Braithwaite et al., 2016 advocated, the ability to learn from crises and implement the necessary organizational structure adjustment is essential to countenance any future crisis. This is a continuous learning approach, where organizations use their experiences from crises to improve their existing strategies and procedures.

To our knowledge, this is the first literature review comprehensively investigating organizational resilience, its measurements, associated factors, and impacts on healthcare workers during the COVID-19 pandemic. Our review has several limitations. First, we relied solely on data from four databases and did not incorporate other potentially relevant sources such as Google Scholar, introducing a risk of selection bias and potentially overlooking valuable research and perspectives. Second, language restrictions further limited our review, as we only considered studies published in English, potentially excluding valuable research published in other languages. Finally, the nature of the data may have hindered our ability to generalize findings accurately, as generalizations based on heterogeneous data can oversimplify complex phenomena and lead to erroneous conclusions.

Researchers need to acknowledge that organizational resilience in public health research is a multi-domain concept that should be understood and operationalized at multiple levels, especially in the context of the specific organizational structure and dynamics of healthcare facilities. For example, organizational agility is a pivotal variable in forthcoming endeavors to enhance crisis management capabilities and organizational resilience. Second, future studies are needed to develop and validate theoretically grounded measurement tools to provide consistent and reliable assessments across different healthcare contexts. Third, more empirical studies (including both qualitative and quantitative research) are needed to identify factors at the community- and policy-level that may affect organizational resilience. Longitudinal studies are also important to capture the evolving nature of resilience over time and during different stages of crises. We also call for intervention and implementation science studies to assess the efficacy of strategies aimed at improving organizational resilience. These studies should evaluate resilience-building efforts and their impact on healthcare providers’ mental health, quality of care, and capacity outcomes at the organizational level in healthcare facilities. Finally, there is a need to explore the role of technology and digital solutions in enhancing organizational resilience. For instance, telehealth use is an innovative implication of advanced technologies in healthcare delivery. The scale-up of telehealth may improve the resilience of healthcare systems on the one hand; the existing “digital divide” and infrastructure disparities may be exacerbated by this implementation on the other hand.

## CONCLUSION

This literature review focuses on the impact of organizational resilience on healthcare workers during the COVID-19 pandemic. Our findings highlight the critical need for building organizational resilience among healthcare workers, particularly through targeted training, psychological support, and adaptable crisis management frameworks. Emphasizing trust in leadership, age-sensitive interventions, and proactive preparedness can significantly enhance organizational resilience, ensuring better mental health outcomes and sustained quality of care during crises. Measurement instruments for organizational resilience vary widely, indicating a need for further theoretical research, conceptualization, and measurement standardization. The limited number of included papers and a lack of intervention studies suggest a large gap between research and practical needs. More empirical studies are needed to further explore factors associated with organizational resilience and inform effective interventions and policies to promote the organizational resilience of healthcare facilities.

## Data Availability

All data produced in the present study are available upon reasonable request to the authors

